# Predicting the efficacy of variant-modified COVID-19 vaccine boosters

**DOI:** 10.1101/2022.08.25.22279237

**Authors:** David S. Khoury, Steffen S. Docken, Kanta Subbarao, Stephen J. Kent, Miles P. Davenport, Deborah Cromer

## Abstract

As a result of the emergence and circulation of antigenically distinct SARS-CoV-2 variants, a number of variant-modified COVID-19 vaccines have been developed. Here we perform a meta-analysis of the available data on neutralisation titres from clinical studies comparing booster vaccination with either the current ancestral-based vaccines or variant-modified vaccines. We then use this to predict the relative efficacies of these booster vaccines under different scenarios.

## Main text

Vaccination provides significant protection from both symptomatic and severe COVID-19. However, the emergence of the antigenically distinct variants such as Omicron and its subvariants has significantly reduced the effectiveness of current vaccine regimes, which are based upon the ancestral (Wuhan-like) variant. This raises two major questions for future vaccine development. Firstly, is there an advantage in switching from the current ancestral-based vaccines to incorporate variant spike proteins? Secondly, if switching to a variant-modified vaccine, how important is it that the immunogen in the variant-modified vaccine is antigenically closely related to the spike protein of the circulating variant? We analysed the data from 8 reports that included a direct comparison of immunogenicity of an ancestral-based vaccine with a variant-modified vaccine^1-8^. They included data from the Sanofi-GlaxoSmithKline, Moderna, and Pfizer-BioNTech vaccine constructs incorporating the Beta, Delta, or Omicron BA.1 spike proteins (either alone or in combination with each other or the ancestral variant spike proteins). We then used a validated model relating neutralisation titres to vaccine effectiveness to estimate the changes in vaccine protection under different booster regimens^9-11^.

To compare the average magnitude of boosting between ancestral-based vaccines and variant-modified vaccines, we first compared the rise in neutralisation titre between pre-booster and post-booster titres. Considering neutralisation for all variants reported in the studies, we found that an ancestral-based vaccine increased neutralisation titres by a mean of 11-fold from pre-booster titres (95%CI 8-15.2) (Fig 1A). Although variant-modified vaccines did not show an improvement in neutralisation towards ancestral SARS-CoV-2 compared to the ancestral-based vaccines (p= 0.46), the variant-modified vaccines produced more potent neutralisation of the variants tested (Fig 1B). Considering only neutralisation of SARS-CoV-2 variant strains, we found that variant-modified vaccines on average produced 1.51-fold [95% CI 1.4-1.6] higher titres than the equivalent ancestral-based vaccine (p<0.0001, Fig 1B). To determine if this superiority of the variant-modified vaccines was only true when the variant-modified vaccine matched the strain used in the neutralisation assay, we compared the level of boosting to the homologous strains (ie: same variant in vaccine and neutralisation assay) versus non-homologous strains (neutralisation of variants not included in the vaccine). Boosting was slightly higher against homologous antigens (1.75 vs. 1.31-fold, p=0.00032). We found no significant differences when stratifying results for monovalent versus bivalent vaccines (p = 0.73, see Fig S1).

**Figure 1.**
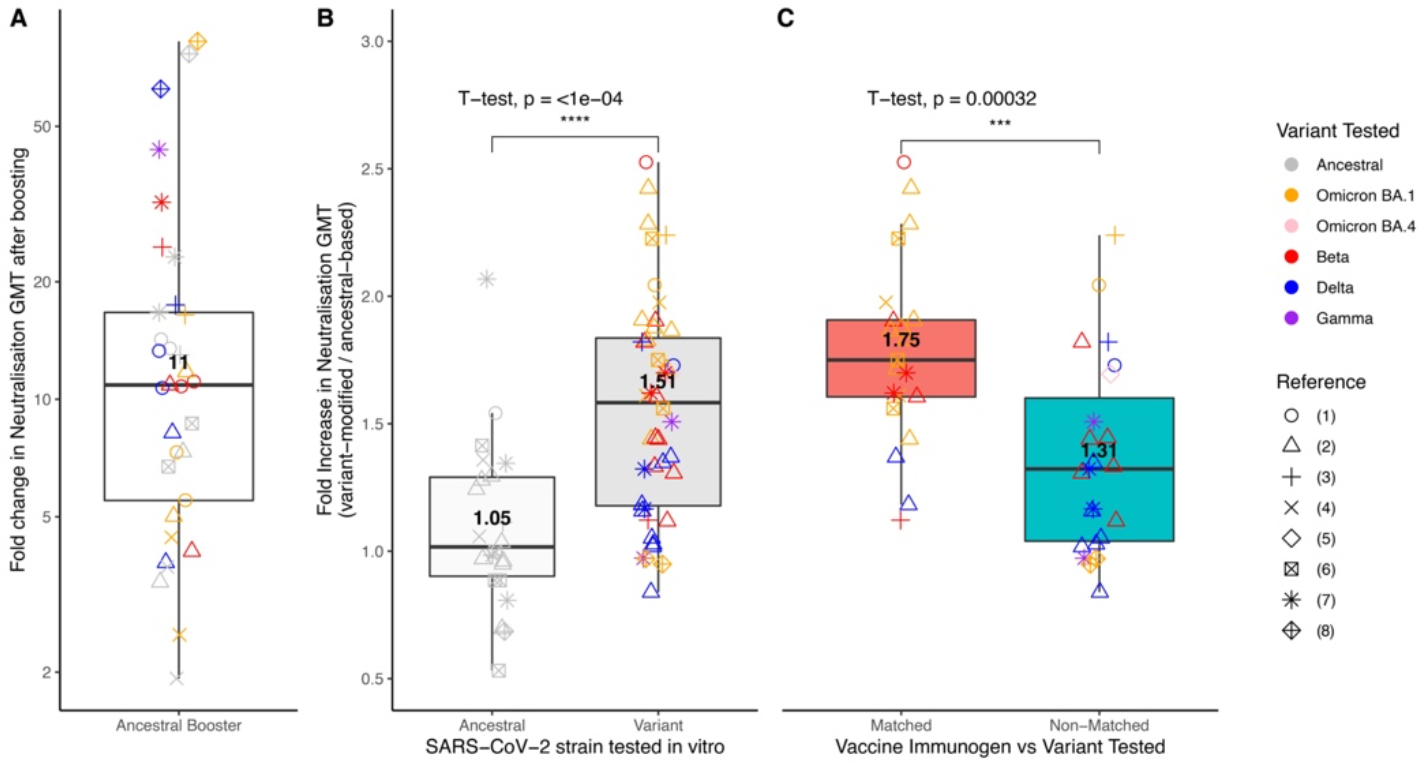
Aggregated neutralisation data on boosting with both ancestral-based and variant-modified vaccines. (A) Fold change in neutralisation titres after boosting with an ancestral-based vaccine. Change in titres against different tested variants are depicted in different colours. (B) Improvement in neutralisation titres (shown as fold increase) when boosting with a variant-modified vaccine compared to an ancestral-based vaccine. Improvement is shown when testing against the ancestral SARS-CoV-2 variant in-vitro (left), and when testing against other variants in-vitro (right). (C) Fold increase in neutralisation titres after boosting with a variant-modified vaccine compared to an ancestral-based vaccine depending on whether the variant tested in-vitro matched the vaccine immunogen (red, left) or did not match (blue, right) the vaccine immunogen. For panels B and C, t-tests were performed on the log10-transformed values.

To estimate the clinical benefits of the 1.5-fold improvement resulting from switching from ancestral-based boosters to variant-modified boosters, we used a model that correlates neutralisation titres with observed clinical protection^9^. This model was originally parameterised from phase 3 clinical trials of seven vaccines, and has subsequently been validated for the beta, delta and omicron variants^10,11^, as well as showing good agreement with individual-based studies of protective immunity^12^. Here we use the model to predict the relative benefits of booster vaccination with an ancestral-based vaccine compared to a variant-modified vaccine. We considered the relative impact of an ancestral-based booster that delivers an 11-fold boost in neutralising antibody titres and a variant-modified booster delivering a further 1.5-fold higher titre than the ancestral booster (i.e. a 16.6-fold increase in neutralising antibody titres compared to pre-boost, consistent with the analysis above). We estimated the average protection provided by the different boosters over a six month period, assuming antibody titres decay at the same rate for both vaccines (with a half-life of 108 days^9,10,13^).

The relative benefits of a variant-modified vaccine are very dependent on the underlying (pre-booster) population immunity to infection for the currently circulating variant (Fig 2). If we consider a previously vaccinated or infected population that already has 50% protection from symptomatic infection (compared with a naïve and unvaccinated population), we find that an ancestral-based booster giving an 11-fold boost in neutralising antibody titres would increase the average protection over a six month period against symptomatic infection from 50% to 85.6%, while a variant-modified booster that is 1.5-fold more potent would provide an average of 90.2% protection (i.e. the variant-modified vaccine results in an overall average improvement in protection of 4.6 percentage points compared to the ancestral-based vaccine, Fig 2A and C). Similar analyses can be used to predict comparative vaccine effectiveness against severe COVID-19 under the same assumption. A population with 50% protection from symptomatic infection is predicted to have 86.6% protection from severe COVID-19^9,11^. Boosting with an ancestral-based booster that increases neutralising antibody titres by 11-fold is expected to increase this to an average of 98% protection from severe disease over the six month period following boosting, and a variant-modified booster producing 1.5-fold higher neutralising antibody titres would increase protection to an average of 98.8% (i.e. an additional 0.8 percentage points of protection on average from severe COVID-19 compared to an ancestral-based booster, Fig 2B and D). This would correspond to 8 additional severe cases averted for every 1,000 severe cases that would have occurred in a naive population over the six month period. However, in cases where the underlying immunity to the circulating variant is higher or lower (such as might occur in the context of an antigenically distinct variant or after waning of immunity) the relative benefits of a variant booster compared to an ancestral booster can vary. In general, the lower the pre-booster immunity the greater the relative benefit of a variant-modified booster compared to an ancestral-based booster, peaking with around 10 percentage points additional protection from symptomatic infection when the population has only 11% pre-existing protection against symptomatic disease (Fig 2D).

**Figure 2.**
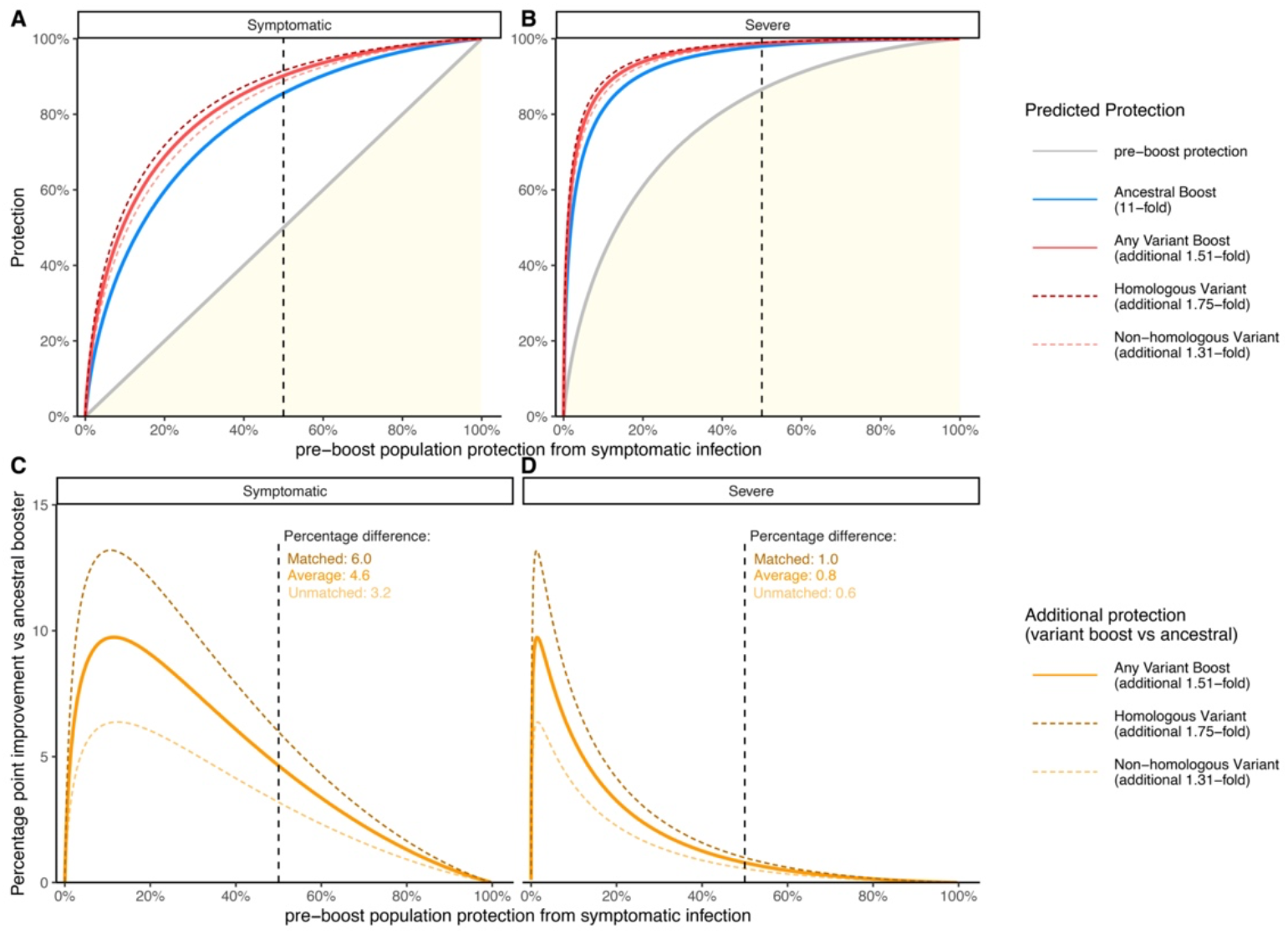
Estimated improved protection from a variant-modified booster over an ancestral-based booster. (A and B) Levels of protection against symptomatic (A) and severe (B) COVID-19 after either no boost (grey), an ancestral-based boost (blue) or a variant-modified boost (red) for varying levels of pre-boost population protection from symptomatic protection. Solid red line shows protection for any variant-modified vaccine, dashed red lines show protection for a matched variant-modified vaccine (dark red) and non-matched variant-modified vaccine (light red). (C and D) Average improvement in protection from symptomatic (C) and severe (D) COVID-19 for a variant-modified vaccine over an ancestral-based vaccine over the six months following boosting. Solid line shows protection for any variant-modified vaccine, dashed lines show protection for a matched variant-modified vaccine (dark orange) and non-matched variant-modified vaccine (light orange). Text at the top of panel C and D indicate the percentage point improvement in protection over an ancestral-based booster when the pre-boost population protection against symptomatic disease is 50%.

The analysis above considers the benefit of any variant-modified booster (regardless of whether or not it is antigenically similar to the circulating variant). Given that the relative advantage of variant modified boosters was found to be higher to homologous than non-homologous variants (1.75-fold vs 1.31-fold), using the same approach we also estimated the relative advantage of a booster that did or did not match the antigenicity of the circulating variant over the first six months after boosting. We found a maximum benefit of a 13 percentage point improvement in protection for boosters against antigenically related strains and 6 percentage point improvement for antigenically distinct boosters (Fig 2C and D). However, the trajectory of SARS-CoV-2 variant evolution is unclear and the frequency with which vaccine composition will be updated is not known.

This approach provides a method for predicting the comparative effectiveness of different booster vaccine formulations, informed by relative improvements in neutralisation titres and changes in titres against different variants. Synthesis of the currently available data suggest that variant-modified booster vaccination can provide significantly higher (1.5-fold) neutralisation titres to a diversity of current and historic SARS-CoV-2 variants compared to ancestral-based boosters. This is predicted to provide up to a maximum of a 9.7 percentage point increase in protection (dependent on the pre-boost level of population protection). This analysis includes a number of important caveats. Firstly, it analyses a limited number of studies of variant-modified boosters, which were carried out largely by vaccine manufacturers^1-8^. Secondly, these studies reported neutralisation titres against a limited set of variants that were not the same across studies. In addition, it relies on predicting vaccine efficacy from neutralisation titres. Although this approach has been validated in a number of contexts^9-11,14,15^, it cannot replace clinical studies of vaccine effectiveness. Thus, it will be important to validate these predictions in future. Despite these limitations, this work provides a quantitative and evidence-based mechanism to estimate and compare the relative benefits of different vaccine modifications. This suggests that a large proportion of the benefit comes from receiving any booster at all (including an ancestral-based booster). Use of a variant-modified vaccine is expected to provide a modest increase in protection, which may be slightly greater in cases where the vaccine immunogen is more antigenically related to the circulating variant or if immunity has waned. However, even if the SARS-CoV-2 variant circulating at the time of vaccination is relatively antigenically distant from immunogen in the variant-modified booster, an elevated level of protection (when compared to either no booster or an ancestral-based booster) is still expected. Importantly, the overall benefit of variant-modified vaccines will likely be determined by other factors, including the time since vaccination (waning of immunity), relative availability, cost and community acceptance of variant modified vaccines over existing vaccines.

## Data Availability

All data and code used will available with no restrictions following publication

## Supplementary Figures

**Supplementary Figure 1.**
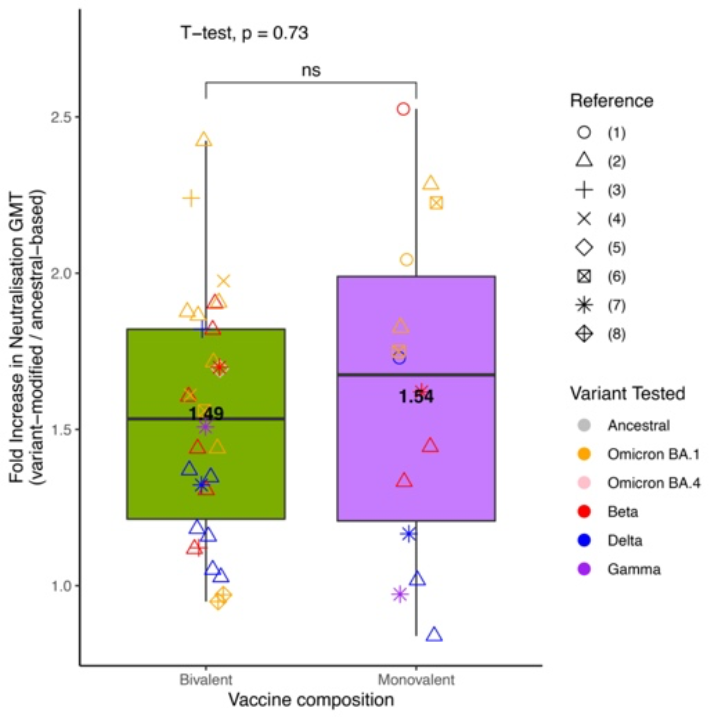
Comparison of the improvement gained for variant-modified boosts compared to ancestral-based boosts split by monovalent and bivalent vaccines. T-test was performed on the log10-transformed values.

## Ethics statement

This work was approved under the UNSW Sydney Human Research Ethics Committee (approval HC200242).

## Authorship Statement

DC, MPD and DSK contributed to study design. DC, DSK and SD performed the data extraction, curation and analysis. DC, DSK, and SD performed the data analysis. DC, MPD, DSK, KS and SJK contributed to shaping the direction of the work. All authors contributed to the writing and reviewed and approved the final report.

## Data Availability Statement

Data and code will be posted on a public repository after publication.

