## Supplementary Materials for "Predicting the efficacy of variant-modified COVID-19 vaccine boosters"

### Supplementary Tables

| Refer-ence | Previous immunological history of study participants | Dosage | Vaccines reported | Variants tested | Neutralisation measured after[^] | Data derived from |
|---|---|---|---|---|---|---|
| 1 | BNT162b2 primary vaccination 3-7 months prior, no prior infections | nr | BNT162b2 (Ancestral)<br>MVD614 (Ancestral)<br>MVB.1.351 (Beta) | Ancestral, Beta, Delta, BA.1 | 15 d | Figure 1 |
| 2 | primary vaccination and booster (~95% received mRNA vaccines for both, remainder received Ad26.COV2.S for one or both of primary / booster doses). Booster given 112-333 days prior<br>Grouped by either (A) no prior infections or (B) infection >=110 days prior | 50µg | mRNA-1273 (Ancestral)<br>mRNA-1273.529[#] (BA.1)<br>combination of mRNA-1273.351 and mRNA-1273.529[#] (Beta+BA.1)<br>combination of mRNA-1273.617.2[#] and mRNA-1273.529[#] (Delta+BA.1),<br>combination of mRNA-1273 and mRNA-1273.529[#] (Ancestral+BA.1) | Ancestral, Beta, Delta, BA.1 | 15 d | Table S4 |
| 3 | mRNA-1273 primary vaccination >6 months prior, no prior infections | 50µg<br>100µg (mRNA-1273.211 only) | mRNA-1273 (Ancestral)<br>mRNA-1273.211 (Ancestral + Beta) | Ancestral, Beta, Delta, BA.1* | 28 d | Table S4 |
| 4 | mRNA-1273 primary vaccination and mRNA-1273 booster >3 months prior. Grouped by either (A) no prior infection and (B) infection >3 months prior | 50µg | mRNA-1273 (Ancestral)<br>mRNA-1273.214 (Ancestral + BA.1) | Ancestral, BA.1, BA.4/5* | 28 d | Tables S7 and S8 |
| 5 | primary vaccination and booster[§], no prior infection | 50µg | mRNA-1273 (Ancestral)<br>mRNA-1273.214 (Ancestral + BA.1) | BA.4/5 | 1 mo | Text |

| | | | | | | |
|---|---|---|---|---|---|---|
| 6 | BNT162b2 primary vaccination and BNT162b2 booster >4.7 months prior, no prior infections | 30µg<br>60µg | BNT162b2 (Ancestral), BNT162b2 Omi (BA.1), BNT162b2 + BNT162b2 Omi (Ancestral + BA.1) | Ancestral, BA.1, BA.4/5* | 1 mo | Slides labelled CC8-CC19 |
| 7 | mRNA-1273 primary vaccination >5 months prior, no prior infection | 50µg<br>20µg (mRNA-1273.351 only) | mRNA-1273 (Ancestral)<br>mRNA-1273.351 (Beta)<br>mRNA-1273.211 (Ancestral + Beta) | Ancestral, Beta, Gamma, Delta* | 15 d or 29 d | Figures 3, 4 and S1 |
| 8 | mRNA-1273 primary vaccination, no prior infections | 100ug<br>50ug for mRNA-1273.211) | mRNA-1273 (Ancestral)<br>mRNA-1273.211 (Ancestral + Beta),<br>mRNA-1273.213 (Beta + Delta) | Ancestral, Beta, Delta, and BA.1* | 29 d | Figure S6 |

*Supplementary Table 1 Reports of Variant modified vaccines used in this analysis*

nr = not reported, BA.1 and BA.4/5 refer to Omicron BA.1 and Omicron BA.4/5 variants respectively, # technical names not listed in reference, but inferred from clinical trial number NCT05289037, * Results were not reported for all booster-variant combinations, ^ d=days, mo=months, § details of primary and booster vaccine not listed

### Identifying Studies to use in this analysis

We surveyed the academic literature (published works and pre-prints), manufacturer press-releases and information provided to regulators for data on variant-modified vaccine immunogenicity. Where we identified multiple reports of the same underlying cohort or trial (e.g. a pre-print and press-release presenting the same information from a specific trial), we collected information from the report containing the most complete information only. We identified 8 reports with unique information on variant-modified vaccine immunogenicity up to 15 July 2022.

The studies identified for this analysis are shown in **Supplementary Table 1**.

### Extracting data from relevant studies

For each study identified in **Supplementary Table 1**, we extracted information on the booster composition(s), the dose(s) administered, and the variants against which neutralising antibodies were tested in vitro. For each vaccine immunogen / dose / in vitro variant combination we recorded the geometric mean neutralising antibody titre (GMT) pre- and post-vaccination (for all reported assays). Where available, neutralising antibody titres at two weeks following the booster dose were used. However if these were not available, titres from up to one month following boost were used.

In the one case where exact neutralising antibody levels were not reported[6] the ratio between neutralising antibody levels against a variant of concern and against the ancestral virus was extracted instead.

For all further analysis, extracted data was grouped by study, vaccine composition, assay and booster dose to ensure comparisons were only made between matched ancestral-based and variant-modified vaccines.

### Fold-rise in antibodies after boosting

To calculate the fold rise in neutralising antibody levels after boosting (with either a variant-modified vaccine or an ancestral-based vaccine) we divided the reported GMT post-boost neutralising antibody levels for each variant tested in vitro with the matched GMT pre-boost neutralising antibody levels for the same in vitro variant (matching data by study, dose and assay). That is, the formula governing the fold-change (*FC*) calculation is

$$FC(v,m,d,s,a) = \frac{NAb_{post}(v,m,d,s,a)}{NAb_{pre}(v,m,d,s,a)}$$

*Equation 1*

Where, $NAb_{post}$ is the GMT of the neutralising antibody titres following booster dose, $NAb_{pre}$ is the GMT of the neutralising antibody titres prior to the booster dose, $v$ is the variant tested, $m$ is the makeup of the booster vaccine, $d$ is the dose administered, $s$ is the study and $a$ is the assay used.

**Fold-rise in neutralising antibody titres for a specific booster vaccine**

For a specific booster vaccine makeup $m$ we can determine the geometric mean fold rise in neutralising antibody titres, $\overline{FC(m)}$, by taking the geometric mean of Equation 1 over all available variants tested, doses administered, studies considered and assays used. Thus, the geometric mean fold rise in neutralising antibody titres for a specific booster vaccine is given by

$$\overline{FC(m)} = \left(\prod_{all\ v,d,s,a} FC(v,m,d,s,a)\right)^{\frac{1}{N_m}}$$

*Equation 2*

Where $N_m$ is the number of unique combinations available in our extracted data of $v, d, s$ and $a$ for vaccine makeup $m$.

**Improvement for a variant-modified vaccine boost compared to an ancestral-based vaccine boost**

To estimate the improvement gained by boosting with a variant-modified vaccine over boosting with an ancestral-based vaccine, we compared the neutralising antibody titres following boosting with a particular variant-modified vaccine to the corresponding neutralising antibody titres after boosting with an ancestral-based vaccine (matching data based on study, variant tested in vitro, assay and dose administered). The formula governing the improvement (*I*) is given by:

$$I(v,m,d,s,a) = \frac{NAb_{post}(v,m,d,s,a)}{NAb_{post}(v,m_A,d,s,a)}$$

*Equation 3*

Where $v, m, d, s$ and $a$ are as defined above, and $m_A$ refers to the ancestral-based vaccine.

We can then calculate the geometric mean improvement for a group $K$ of variant-based boosters, $\overline{I(K)}$, using the formula

$$\overline{I(K)} = \left(\prod_{m \in K}\left(\prod_{all\ v,d,s,a} I(v,m,d,s,a)\right)^{\frac{1}{N_m}}\right)^{\frac{1}{N_K}}$$

*Equation 4*

Where $N_K$ is the size of the group $K$.

**Matching of vaccine immunogen and variant tested**

We determined that the variant tested ($v$) was matched to vaccine immunogen (or vaccine make-up, $m$) if the variant tested was one of *any* of the variants that went into the vaccine makeup. Therefore, when testing neutralisation of the Omicron BA.1 variant in vitro, this would have been considered to be matched with any of the following vaccines; (i) a monovalent BA.1 vaccine, (ii) a bi-valent vaccine containing BA.1 and ancestral virus spike or (iii) a bi-valent vaccine containing BA.1 and any other variant spike (e.g. Beta or Delta). However, it would be considered non-matched to either an ancestral-based vaccine, or a variant modified vaccine that did not include the BA.1 variant spike. By a similar argument, when testing the neutralising antibody titres against the ancestral virus in vitro, the ancestral variant would be considered to be matched with the ancestral-based vaccine, as well as with any variant-modified vaccine that also included the ancestral spike in its make-up.

**Waning of neutralising antibodies**

Following boosting, neutralising antibodies are assumed to wane back to their pre-boost level with a half-life of 108 days [9-11]. Therefore, if the neutralising antibody titres prior to boosting were given by $n_0$, then at $t$ days after an $f$-fold boost in neutralising antibody titres due to vaccination, neutralising antibody titres, $n_{boost}(t, f, n_0)$ would be given by

$$n_{boost}(n_0, f, t) = \max\left(f n_0 e^{-\frac{\ln(2)t}{108}}, n_0\right)$$

*Equation 5*

**Calculating the average efficacy over a period of time, $T$, after boosting**

In order to determine the average efficacy within a group of subjects over a fixed period of time following boosting (either by an ancestral-based or variant-modified booster vaccine) we first need to know the pre-boost level of neutralising antibody titres. This is equivalent to knowing the pre-boost symptomatic efficacy (labelled as $VE_0$) within the population from which the subjects are taken.

Given $VE_0$, the fold rise ($f$) in neutralising antibody titres due to the vaccine boost and a period of time ($T$), we can follow the steps 1-4 below to calculate the average efficacy over the time period after boosting:

1) *Establish the average baseline neutralising antibody titres in the population.*

   We can calculate the average neutralising antibody titres within the population using our established relationship between neutralising antibody levels and vaccine efficacy[10]. This is done by using the inverse of the relationship between neutralising antibody titres and vaccine protection previously established in *Khoury et. al*[10]*.* For neutralising antibody titres given by $n$, the original relationship takes the form

$$VE(n) = \int_{-\infty}^{\infty} N(x, \log10(n), \sigma) \frac{1}{1 - e^{-k(x - x_{50})}} dx$$

*Equation 6*

   Where $VE(n)$ is the vaccine efficacy (compared to a naïve population) for a population with average neutralising antibody titres of $n$, N is the probability density function of a normal distribution with mean $n$ and standard deviation $\sigma$, $\sigma$ is the previously reported standard deviation (0.46) in neutralising antibody titres and $k$ (3.1) and $x_{50}$ ($\log_{10}$(0.20)) are logistic parameters fitted in *Khoury* et. al.[10] from randomized controlled trial data.

   Given a pre-boost level of protection from symptomatic disease of $VE_0$, we can numerically calculate the corresponding average neutralising antibody titres in the population, $n_0$, by using the inverse of Equation 6. We will represent these population average neutralising antibody titres for a given pre-boost symptomatic efficacy, $VE_0$, as $\hat{n}(VE_0)$.

2) *Calculate the neutralising antibody titres at a given time, $t$, after boosting*

   For a given fold-boost in neutralising antibody titres due to a vaccine, we use $n_0$ determined in step 1 above and Equation 5 to calculate the neutralising antibody titres $t$ days after boosting.

3) *Establish the new efficacy (compared to a naïve population) at a given time, $t$, after boosting*

   For the neutralising antibody titres calculated in step 3 above, we use Equation 6 and Equation 5 to calculate the new efficacy $t$ days after boosting. Therefore, efficacy at a given time, $t$, after an $f$-fold boost in a population with average neutralising antibody titres of $n_0$, $VE_{boost}(n_0, f, t)$, is given by:

$$VE_{boost}(n_0, f, t) = \int_{-\infty}^{\infty} N(x, log10(n_{boost}(n_0, f, t)), \sigma) \frac{1}{1 - e^{-k(x - x_{50})}} dx$$

*Equation 7*

4) *Determine the average efficacy over time-period $T$ after boosting*

   We next integrate Equation 7 from the time of boosting until the end of the time-period $T$ to determine the average efficacy over time-period $T$ after boosting with an and $f$-fold boost in a population with average neutralising antibody titres of $n_0$, $\overline{VE_{boost}}(n_0, f, T)$. This results in the following expression:

$$\overline{VE_{boost}}(n_0, f, T) = \frac{1}{T} \int_0^T VE_{boost}(n_0, f, t) dt$$

*Equation 8*

**Average efficacy improvement for a variant-modified vaccine over an ancestral-based vaccine**

To determine the efficacy improvement for a variant-modified vaccine over an ancestral-based vaccine we first determine the fold-change in neutralising antibody titres for the ancestral-based vaccine, $\overline{FC(m_A)}$ using Equation 2 and label this as $f_A$. The improvement for a group $K$, of variant-modified vaccines is given in Equation 4 by $\overline{I(K)}$.

Therefore, the average efficacy over time-period $T$ after giving an ancestral-based boost in a population with a pre-boost symptomatic efficacy of $VE_0$ is given by $\overline{VE_{boost}}(\hat{n}(VE_0), f_A, T)$ and the average efficacy over time-period $T$ after giving a variant-specific boost with a variant-specific vaccine from group $K$ in a population with a pre-boost symptomatic efficacy of $VE_0$ is given by $\overline{VE_{boost}}\left(\hat{n}(VE_0), f_A\overline{I(K)}, T\right)$. Combining these expressions together, we find that the improvement in protection for a variant-modified vaccine (from a group $K$ of variant-modified vaccines) over an ancestral-based vaccine (denoted by $IP(K)$) is given by:

$$IP(K) = \overline{VE_{boost}}\left(\hat{n}(VE_0), f_A\,\overline{I(K)}, T\right) - \overline{VE_{boost}}(\hat{n}(VE_0), f_A, T)$$

*Equation 9*

We note that the units of $IP(K)$, the improvement in protection, are in percentage points.
